# Projected cumulative lifetime earnings of physicians

**DOI:** 10.1101/2025.08.19.25333972

**Authors:** Nicholas G. Zaorsky, Christine Lin, Kyle Mani, Daniel E. Spratt, James M. Dahle, Ming Wang

**Author notes:** Corresponding author: Nicholas G. Zaorsky, MD MS Department of Radiation Oncology, University Hospitals Seidman Cancer Center 1100 Euclid Avenue, Cleveland, OH 44106.

## Abstract

**Purpose:** The estimated lifetime earnings of physicians are multifactorial and have not been extensively modeled. This study projects and compares lifetime earnings of US-based physicians across different career pathways.

**Methods:** We developed a customizable model to estimate physician lifetime earnings using data from the publicly available 2016-17 Association of American Medical Colleges (AAMC) and Medical Group Management Association (MGMA) survey results across variable undergraduate training time, MD/DO vs MD-PhD degree programs, attending practice settings, state income tax levels during attending practice, loan repayment options, and promotion timeline. Post-tax earnings were calculated using U.S. tax codes and specialty-specific salary data. We created an interactive web-based interactive application using R Shiny to simulate individualized financial projections. App: https://physician-lifetime-earnings.shinyapps.io/physician-lifetime-earnings/.

**Results:** The median cumulative lifetime earnings averaged across 19 specialties was $6,509,474. Specifically, the average was $5,167,507 across primary care specialties (family medicine, internal medicine, and pediatrics) and $7,696,166 across all specialties excluding primary care. Pursuing two paid undergraduate gap years (vs zero) resulted in a weighted average 5.2% (IQR: 5.2, 5.7) decrease in cumulative lifetime earnings, private practice (vs academia) resulted in a 14.7% (IQR: 8.9, 22.4) increase, and repaying loans at 30% of income (vs obtaining Public Service Loan Forgiveness) resulted in a 6.1% (IQR: 3.0, 6.5) decrease. Completing one undergraduate gap year (vs zero), a combined MD-PhD degree (vs MD/DO alone), and accelerated academic promotion at eight years to full professor (vs 14) all had minimal impact on cumulative lifetime earnings (< 3%).

**Conclusion:** We created a model to compare the lifetime earnings of US-based physicians across different career pathways.

## INTRODUCTION

The lifetime cumulative earnings of physicians have not been recently reported. A study using data from ∼20 years ago identified a strong disparity in lifetime earnings across specialties, with primary care specialties’ lifetime earnings approximately $1 – 3 million less than other specialties.^1,2^ More recent analyses evaluate differences between adults and pediatric specialties^3^, sex differences in physician researcher salaries^4^, physician salaries in US public medical schools^5^, or focus exclusively on subgroups of physicians.^6,7^

Factors that affect lifetime earnings include taking a gap year, completing an MD-PhD degree^8^, specialty choice, promotion timeline in academia, geographical locations, institutions, and post-residency practice settings^9,10^. About 47% of applicants take one to two gap years to affirm interests in medicine, complete prerequisite courses, or prepare for the Medical College Admission Test, but with opportunity costs.^11^ In 2022, the median student debt was $200,000 and can impact specialty choice.^12,13^ Practice settings can have varying responsibilities, compensation, and career satisfaction.^9,10^ Promotion timelines typically require a median of 6-9 years for promotion to associate professor and an additional 7-13 years to full professor but can vary by sex^14,15^ and race.^16^ Loan repayment includes standard repayment plans and the Public Service Loan Forgiveness (PSLF) program. Additional decisions including loan, interest rate, and fellowship training^17,18^ impact long-term physician earnings.

There is no current model that accounts for all these variables. Thus, the objective of this work is to create a comprehensive analysis and interactive model to estimate lifetime earnings of US physicians. In this study, we used two primary data sources: the Medical Group Management Association (MGMA) survey and the Association of American Medical Colleges (AAMC) faculty salary reports. The MGMA DataDive aggregates self-reported salary and bonus data from over 1,000 U.S. medical groups annually, while the AAMC survey compiles faculty compensation from accredited U.S. medical schools, stratified by academic rank and specialty. The results of this work may help physicians with evaluating the financial impact of career decisions.

## METHODS

### Data sources and acquisition

In this study, we utilized salary data from the 2016 MGMA provider compensation reports and the 2017 AAMC reports. Specialty-level head counts were drawn from the AAMC’s Physician Specialty Data Report, which utilizes the AMA Physician Masterfile and state licensure records to compile yearly totals. We used the 2017 AAMC report to align with our compensation percentiles and relied on the most recent (2021) AAMC report solely to update the total number of practicing physicians per specialty; no other AAMC data elements (e.g., compensation) were sourced from 2021.

For academic promotion levels (assistant, associate, full professor), we rely on physician-reported data from these MGMA and AAMC sources and do not estimate these values ourselves. Consequently, the compensation data for each academic rank reflects real-world reporting rather than modeled assumptions. By contrast, in specialties where specific academic rank salary data were missing (e.g., pediatric otorhinolaryngology), we assumed a 10% salary increase per promotion to fill any gaps.^19^ We have now also created a customizable option in the web tool that allows users to override the default promotion-step increase, so that users may enter their own %-increase values for each promotion level directly in the Shiny interface.

### Model Design and Assumptions

We developed a customizable model to estimate yearly, and cumulative post-tax lifetime earnings based on five key parameters: (I) undergraduate education, (II) medical school, (III) graduate medical education (GME), (IV) employment, and (V) loan repayment. The model incorporates variables across these domains to provide individuals with a comprehensive tool for financial planning based on their specific educational and career trajectories.

In designing the model, we limited the inputs to five parameters because previous literature has demonstrated these to be determinants of physician lifetime earnings. First, the timing of undergraduate education (including one-or two-year gap periods) directly shifts entry into residency. Economic models have previously shown that each gap year reduces lifetime after-tax earnings by roughly 2–5 percent because of foregone attending-level salary.^20^ Second, the medical-school pathway (MD/DO vs MD-PhD) changes both tuition burden and total training time. Despite funded tuition, recent analyses demonstrate that MD-PhD graduates accrue slightly lower career earnings than MD peers in the same specialty.^21^ Third, GME specialty and duration set the starting post-training salary and therefore appear as standard covariates in physician-economics studies. Fourth, employment setting introduces well-documented compensation differentials as large national surveys consistently report a 10–15 percent salary premium for private-practice physicians over academic colleagues, independent of region or gender.^22^ Finally, loan-repayment strategy strongly shapes early-career cash flow. In fact, Public Service Loan Forgiveness (PSLF) enrolment has tripled among new physicians in recent years,^23^ and multiple educational-debt studies link repayment burden to specialty choice and long-term earning power.^24^

For the undergraduate parameter (I), the model allows individuals to input their starting age, annual costs not covered by loans, annual loan amounts with interest rates, and the number of gap years taken. If gap years are included, the model adjusts for annual costs not paid by loans and annual income during that time. Similarly, for medical school (II), users specify their degree type (MD, DO, or MD-PhD), annual medical school loan amounts with interest rates, and any gap years, factoring in associated costs and income.

The GME parameter (III) includes the option to select from 98 specialties recognized by the AAMC, accounting for the number of years of training and state income tax rates. For employment (IV), the model offers multiple practice types, including academic, private practice, hospital-employed, and pure physician-scientist. It incorporates variables such as retirement age and estimated income percentile. We calculated salaries by decile (e.g., 10th, 20th, 30th, 40th) and included this parameter in the model’s user interface. This baseline scenario merely applies a constant percentile, and users can customize percentiles using the source code as desired. For hospital-employed physicians, we assumed an annual salary equivalent to that of an associate professor, while for pure physician-scientists, we assumed an annual salary equivalent to the National Institutes of Health (NIH) cap of $200,000. Users can also adjust their time on the academic track and select years until achieving partnership for private practice. For private practice, the model assumes a pre-partnership track salary at the 30th percentile and post-partnership track salary at the 50th percentile as the reference level, with options to adjust salaries based on the 25th and 75th percentiles.

Finally, the loan repayment (V) parameter provides options of no repayment, Public Service Loan Forgiveness (PSLF), or repayment out of pocket. Post-tax yearly and cumulative lifetime earnings are calculated based on U.S. tax codes, incorporating wide income ranges using median salaries for each specialty and weighted averages based on the proportion of physicians in each specialty, as reported in the 2021 AAMC survey.

We have extended our tax-engine so that all lifetime-earnings calculations, and the standalone marginal/effective-tax panel, draw from a single “Filing Status” selector offering the three standard U.S. statuses: Single, Married Filing Jointly, and Head of Household. Federal income tax is now computed via a fully vectorized routine against the 2017 IRS marginal-rate tables for the chosen status, while state income tax remains a user-adjustable flat rate (default 5 %). We retain a simple, transparent approach by excluding itemized deductions, dependents, retirement-account deferrals, and local taxes. Every change to filing status or state rate immediately updates both pre-tax and post-tax cash-flow streams.

The reference levels for our model are derived from national averages and aim to provide a starting point for users to adapt to their personal circumstances. The reference levels for our model include four years of undergraduate education with an annual tuition of $40,000, an MD/DO degree with annual loans of $55,000 at 8% interest, no gap years at any point in training, a flat state income tax of 5%, practicing at an academic center on a tenure track with seven years as an assistant professor and seven years as an associate professor before promotion to full professor, successfully obtaining PSLF, and retiring at age 65. These values are based on national averages. Using R and RShiny, we translated our model into an interactive web-based application (App: https://physician-lifetime-earnings.shinyapps.io/physician-lifetime-earnings/), enabling users to customize inputs for each parameter to generate individualized financial projections (R Core Team, 2022).^25^

### Probabilistic Sensitivity Analysis

To characterize the impact of sampling uncertainty in the published MGMA/AAMC percentile tables, we performed a Monte-Carlo probabilistic sensitivity analysis (PSA). For every specialty and academic or private-practice rank we fitted a log-normal distribution whose location parameter μ and the scale parameter σ reproduce the reported 25th, 50th, and 75^th^ percentiles. We then drew 5 000 salary sets per specialty, generating synthetic compensation tables that span the full range of ranks. Each table was passed through the deterministic lifetime-earnings engine under baseline assumptions (age-18 matriculation, MD degree, no gap years, academic career track with standard promotion timelines, retirement at age 65). Federal (2017 single-filer) and state (5 %) income taxes, along with 10 % loan repayment during post-graduate years (consistent with Public Service Loan Forgiveness expectations), were applied exactly as in the primary analysis. The resulting distribution of 5 000 cumulative, post-tax lifetime totals per specialty is summarized by the median and 95 % confidence interval (2.5th–97.5th percentiles).

## RESULTS

We found that the median cumulative lifetime earnings widely range for physicians in different specialties (Figure 1). The weighted average of median cumulative lifetime earnings across all specialties for physicians was $6,509,474. Specifically, for primary care physicians (i.e., in general internal medicine, family medicine, and general pediatrics), the net lifetime earnings were $5,167,507. For non-primary care physicians (i.e., all others), the net lifetime earnings were $7,696,166 (Figure 2).

**Figure 1:**
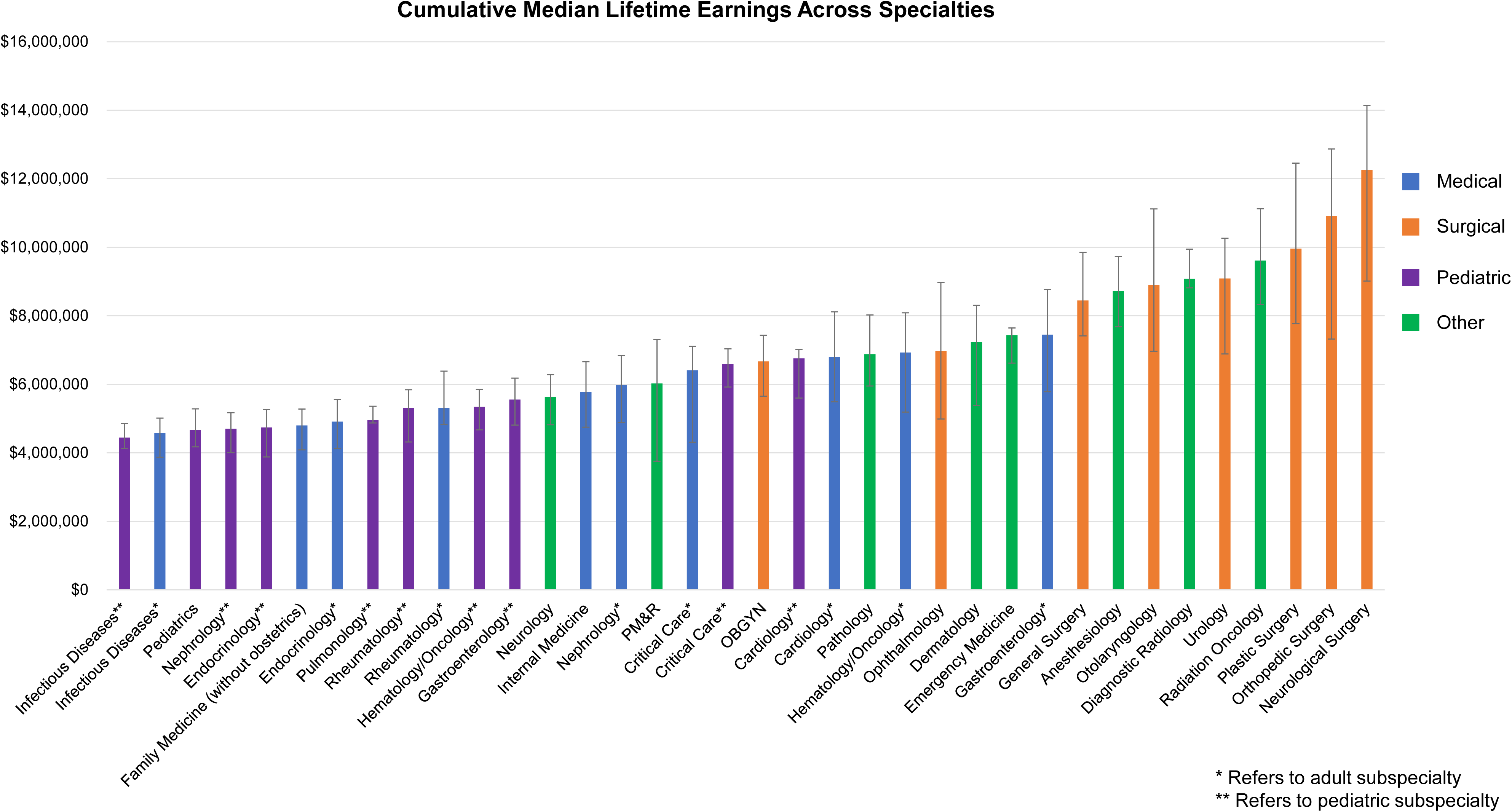
Median cumulative lifetime earnings for each specialty. Modeled cumulative physician lifetime earnings across all specialties, assuming reference level for all variables included: no gap years, median amount of student loans, MD degree, practice setting in an academic center with promotion to full professorship requiring 14 years, flat state income tax of 5% during attending practice, and achieving public service loan forgiveness. Each bar represents the cumulative earnings assuming 50^th^ percentile salary. The lower and upper error bars assume the 25^th^ and 75^th^ percentile salaries respectively.

**Figure 2:**
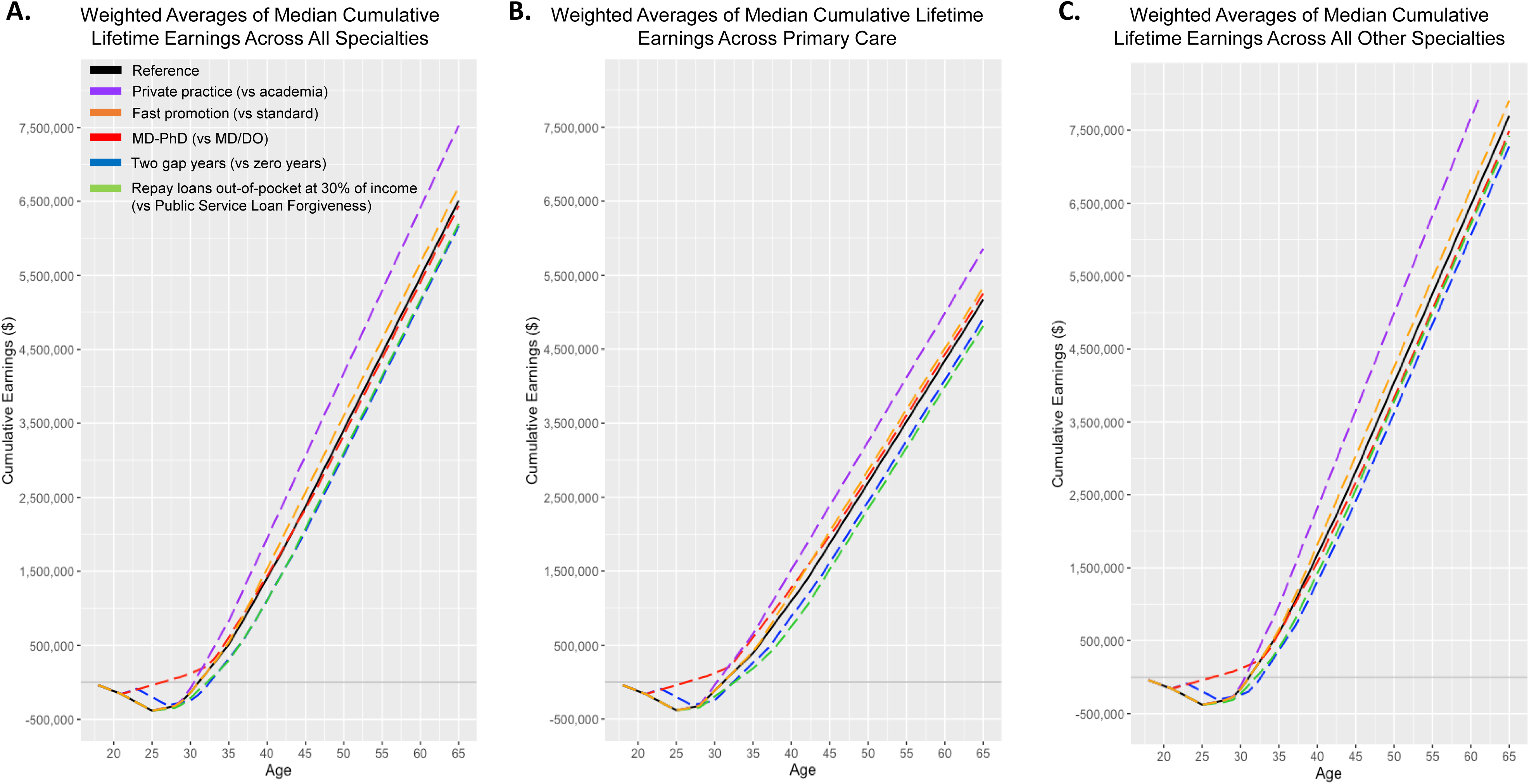
Median cumulative lifetime earnings averaged over specialties over time. (A) Weighted mean of cumulative lifetime earnings across 19 selected specialties. The black solid line represents the reference level across all categories. The purple dashed line represents private practice after residency (vs academia), the orange line represents an accelerated timeline with four years for promotion to associate professor and four years for promotion to full professor (vs seven years each), red line represents MD-PhD degree (vs MD), green line represents repaying loans at 30% out-of-pocket (vs obtaining Public Service Loan Forgiveness), and blue line representing taking two gap years (vs zero gap years). (B) Weighted mean of cumulative lifetime earnings across primary care (family medicine, internal medicine, and pediatrics). (C) Weighted mean of cumulative lifetime earnings across all specialties other than primary care.

Delaying medical education through gap years prior to medical school had an overall negative impact on cumulative lifetime earnings across all specialties, with a 2.6% (IQR: 2.6, 2.85) decrease for those taking one gap year and a 5.2% (IQR: 5.2, 5.7) decrease for two gap years (Figure 3). This decrease in earnings was observed in both primary care fields and non-primary care fields (Figure 2). Supplemental Table 2 shows the break-even age for each of the scenarios depicted in Figure 2. While there was a slight advantage in earnings during the early to mid-twenties from pursuing one to two paid gap years versus zero gap years, this difference was quickly attenuated by the late-twenties, and ultimately had a negative impact thereafter and delayed achieving positive net earnings in contrast to those who did not take gap years. (Figure 3).

**Figure 3.**
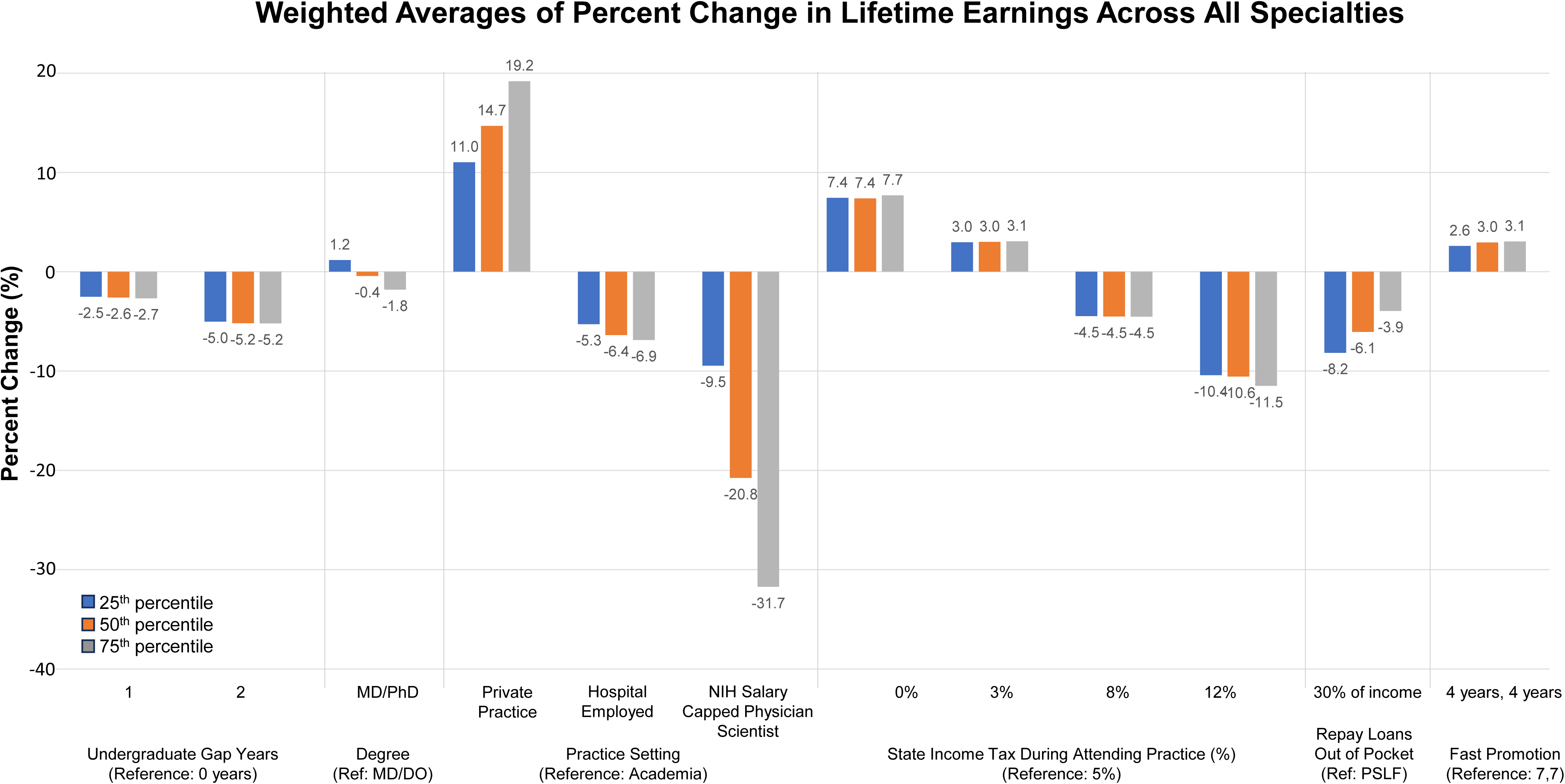
Weighted average of percent change in lifetime earnings. Weighted averages of percent change in lifetime earnings assuming a reference level of no undergraduate gap years, MD degree, academic practice setting, 5% state flat state income tax during attending practice, and pursuing Public Service Loan Forgiveness. Additional assumptions include a four-year undergraduate degree with an annual tuition of $40,000, pursuing an MD degree with annual loans of $55,000 at 8% interest, and retiring at age 65. Values averaged across anesthesiology, dermatology, emergency medicine, family medicine, internal medicine, neurology, obstetrics-gynecology, ophthalmology, orthopedic surgery, otolaryngology, pathology, pediatrics, physical medicine and rehabilitation, psychiatry, radiation oncology, diagnostic radiology, general surgery, neurosurgery, and urology.

Completing a combined MD-PhD program resulted in an overall negligible impact of a 0.43% decrease on cumulative lifetime earnings across all specialties (Figure 3). However, specifically for primary care physicians, there was a minimal 2.3% increase in cumulative earnings, while for non-primary care physicians there was a slight 2.4% decrease in cumulative earnings. Those pursuing funded MD-PhD degree programs achieved net zero earnings earlier than those pursuing MD/DO degrees with student loans (Figure 3).

Repaying loans out-of-pocket at 30% of resident and attending incomes resulted in a 6.1% (IQR: 3.03,6.45) decrease in lifetime earnings compared to those who successfully obtained Public Service Loan Forgiveness (Figure 3). The magnitude of this loss ranged from an 11% decrease for pediatricians to a minimal 1.3% decrease for orthopedic surgeons. Overall, in contrast to those who obtained Public Service Loan Forgiveness, those who elected to repay loans out of pocket broke even later, but before their mid-thirties (Figure 2).

Fast promotion (4 years from assistant professor to associate, and then 4 years to full professor) versus median of ∼7 and ∼7 years had an overall slight positive 3.0% effect across all specialties (Figure 3). This was more pronounced for pathologists with a 5.8% increase and otolaryngologists with a 4.9% increase. There was no difference in cumulative earnings for academic dermatologists who were promoted through an accelerated versus standard timeline. Those on faster promotion timelines break even at the same time as the reference group, but experienced higher financial gains later in their careers (Figure 2). Physicians with median amounts of student loans break a net worth of 0 by their mid-30s and achieve cumulative earnings of over $6.5 million (2016 – 2017 US dollars) on average.

The strongest impact on lifetime earnings was the practice setting, with an average 15% (IQR: 8.9, 22.4) increase for those who work in private practice and a 21% (IQR: 18.1, 48.0) decrease for those who pursued a pure physician-scientist career (Figure 3), relative to those who worked in academia. Percent increase in cumulative earnings for those who practice in private settings vs academia also ranged from 4.3% for emergency medicine physicians to 32% for neurologists. Private practice primary care physicians earned up to $5,853,453, while private practice non-primary care physicians earned up to $9,007,675. Physicians in private practice also break even earlier than those in academia (Figure 2). The percent decrease in cumulative earnings for pure physician scientists compared to academia was greatest for neurosurgeons at 63%. However, pursuing a pure physician-scientist career had the opposite effect for family medicine physicians and pediatricians, with a minimal increase in cumulative earnings of 1.1% and 4.2%, respectively.

Across 5,000 Monte-Carlo draws per specialty the sampling uncertainty embedded in the MGMA/AAMC percentile tables produced only modest shifts in projected earnings. Median lifetime totals changed by 0.0 % to +8.9 % relative to the deterministic base case, and no specialty moved more than one rank in the overall earnings hierarchy. The largest upward deviations were seen in Orthopedic (Nonsurgical) (+8.9 %), Ophthalmology (+4.5 %), and Orthopedic: Hip & Joint (+4.3 %), whereas specialties such as Pediatrics: Pulmonology and Radiology: Diagnostic exhibited virtually no change (≤0.2 %), indicating high robustness of their deterministic estimates. Importantly, all credible intervals overlapped the deterministic point estimates and preserved the specialty-ordering observed in the base model (Figure 4). Full numerical results, sorted by absolute percentage change, are provided in Supplemental Table 3, which confirms that sampling variability in the source surveys does not materially alter the policy-relevant conclusions of this study.

**Figure 4.**
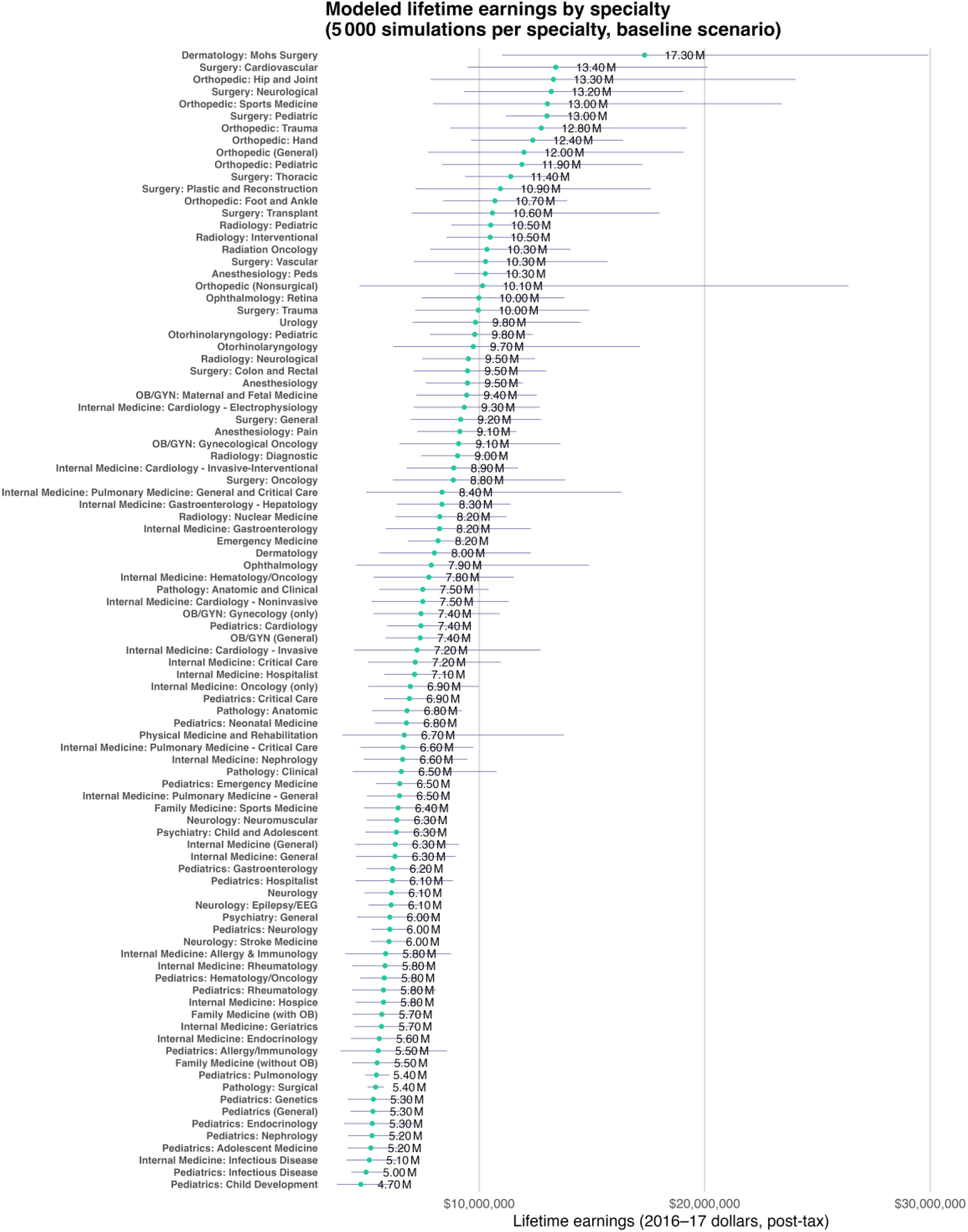
Modeled lifetime earnings by specialty. Median post-tax, cumulative lifetime earnings across 5,000 Monte Carlo simulations per specialty under baseline assumptions (age 18 start, MD degree, no gap years, academic career track, standard promotion timelines, retirement at age 65). Horizontal lines represent 95% credible intervals (2.5th–97.5th percentiles). Pre-attending salaries were adjusted for taxation and 10% loan repayment, aligned with Public Service Loan Forgiveness expectations.

## DISCUSSION

Estimating the cumulative lifetime earnings of a physician is challenging given multiple combinations of contributory factors. We created a model and an accompanying interactive web-based application to estimate yearly and cumulative lifetime earnings for US-based physicians. We found large variations in earnings across specialties, with a difference of $2.5 million between primary care versus non-primary care specialties ($7.7 vs $5.2 million). Our work demonstrates minimal financial impact of pursuing pre-medical gap years and lengthening training time through combined MD-PhD programs. The long-term tradeoff between additional training time and educational debt can be more clearly quantified with this model.

Prior work suggests that the difference in cumulative earnings for specialists vs generalists is due to payment advantages for procedures over office visits.^26^ In our work, post-residency practice setting had the largest effect on cumulative lifetime earnings, with private practice having the strongest positive effect, and pure physician-scientist as the strongest negative effect. Compared to academic physicians, pure physician-scientists with an NIH salary cap earn 20.8% less, which can discourage students and physicians from pursuing research-heavy careers.

Nearly half (49%) of graduating medical students plan to enter loan-forgiveness programs, 83.9% of whom plan to participate in the Department of Education Public Service Loan Forgiveness program. Among family medicine physicians, the rate of PSLF uptake increased from 7% in 2016 to 22% in 2020.^27^ A multi-institutional retrospective study of trainees demonstrated those in a surgical specialty were more likely to pursue PSLF versus those in medical specialties, which may be related to the generally longer length of surgical training.^28^ Loan forgiveness programs help physicians achieve financial independence at a critical time, in their 30s when they may be starting to have a family.

Our approach differs substantially from other recent analyses of physician earnings, such as the Census Working Paper by Gottlieb et al. While the Census study uses microdata to examine broader labor market trends and wage dynamics among healthcare professionals, our study presents a focused, deterministic model that incorporates physician-specific variables (e.g., gap years, MD vs. MD-PhD pathways, academic rank, and loan repayment strategies). In contrast to the macro-level perspective of the Census Working Paper, which addresses overarching wage patterns across various healthcare sectors, our model captures detailed, individualized trajectories for physicians and highlights the financial impact of specific career decisions throughout training and practice. By leveraging specialty-specific AAMC and MGMA compensation data and customizable parameters, our analysis allows trainees and physicians to quantify and compare nuanced outcomes—such as accelerated promotions, loan forgiveness, and private practice vs. academic careers—under different assumptions. In doing so, our work complements large-scale labor market studies, offering targeted insight into the multifaceted pathways of physician earnings and career development.

Our model uses a single time slice of salary data (2016–2017) and tax policy assumptions relevant to that era. Because physician salaries can rise or fall with market forces, inflation, and shifts in reimbursement structures (RVUs), and because policies—such as the NIH salary cap and PSLF requirements—can likewise change significantly, our historical compensation data may not precisely project future earnings. In particular, the NIH salary cap has increased from ∼$195,000 to ∼$240,000 in recent years, and hospitals or universities may impose their own salary caps for research-intensive roles. Similarly, income tax rates fluctuate with legislative changes, and PSLF eligibility criteria could shift in ways that might either bolster or diminish potential loan-forgiveness benefits. For these reasons, our model should be viewed as a framework for understanding how various career decisions can affect lifetime earnings, rather than an exact real-time calculator of future income. We chose to focus on late 2010s data to provide a consistent snapshot, but the tool remains adaptable if updated inputs become available.

As with all modeling studies, our model cannot include all potential cases. Despite multiple customizable variables to capture the most likely points of decision-making, there are scenarios not accounted for. For example, we assume that attending physicians practice in one setting for their entire careers to simplify our model. Additionally, we assume undergraduate and medical school each require at least four years, and do not include the option to complete either earlier. However, this is uncommon, given the median age of medical school matriculation, and that accelerated MD programs are offered by very few institutions.^29^ Additionally, we use data from 2016 and 2017, and salaries have changed since then.^30^ Some specialties have demonstrated stagnant or declining incomes when adjusting for inflation.^30,31^ We also do not account for time spent in the hospital and other sources of hospital-based revenue (e.g. moonlighting), which vary among specialties.^32,33^ Analyzing cumulative earnings standardized by hours worked can be explored in future research. We do not account for initial net worth, financial support from family, other sources of income, retirement matching from employers, pension plans, investment, or retirement funds, etc. An analysis published in 2023 by National Bureau of Economic Research (NBER) quoted average income for physicians between 40-65 at $350,000, which is significantly higher than averages used in our calculations (around $250-300,000, depending on specialty).^34^ Notably, the NBER uses gross income from IRS tax returns, which include W-2 waged, dividends, capital gains, business income, retirement distributions, and taxable investments (which may all be inflated and come from non-patient related sources). In contrast, our analysis only includes physician reported salaries. We also recognize that the assumption that physician-scientists are capped at the NIH salary of 200,000 is likely not the reality for many physician-scientists. Departments can pay above the NIH cap (through other funding mechanisms), especially if the physician-scientist is bringing in clinical dollars through clinical service. Thus, our model is likely overestimating the lifetime earnings deficit caused by a physician-scientist career.

Our model’s post-tax projections use default 2017 U.S. federal brackets tailored to the selected filing status plus a flat state-rate assumption. We intentionally omit itemized deductions, dependents, health-savings/retirement contributions, and municipal taxes; these factors may modestly shift take-home pay but would uniformly scale cumulative earnings without altering the relative rankings or qualitative comparisons across specialties, gap-year scenarios, or repayment strategies. Users seeking individualized estimates can adjust tax parameters directly in the open-source model.

Lastly, our model could be expanded to simulate scenarios such as aggressive early loan repayment to reduce interest costs or delayed payments for near-term financial flexibility, allowing for tailored guidance on their impact on physicians’ cumulative earnings. Despite these limitations to our model, we still capture the most likely options for users to project their lifetime cumulative earnings, making this a widely accessible application.

## CONCLUSION

We created a model to estimate cumulative lifetime earnings for US-based physicians. We identified the financial impact of decisions in career development from undergraduate to early career years. This model can be used by trainees at all levels to illustrate the financial impact of decisions, such as completing research years before medical school or residency, pursuing MD-PhD vs MD alone, and completing fellowships. Physicians with median amounts of student loans break a net worth of 0 by their mid-30s and achieve cumulative earnings of over $6.5 million (2016 – 2017 US dollars) on average.

## Funding/Support

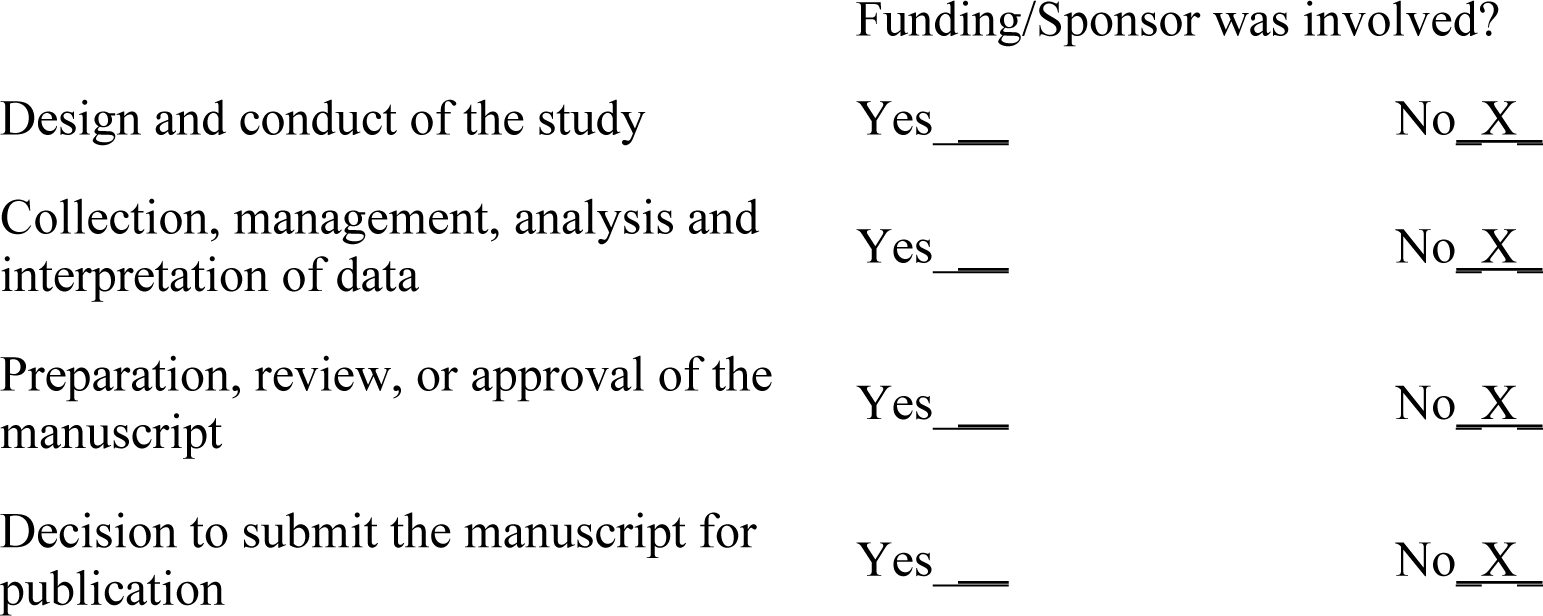

## Data Availability

All data produced in the present work are contained in the manuscript

## SUPPLEMENTAL TABLE

**Supplemental Table 1.**
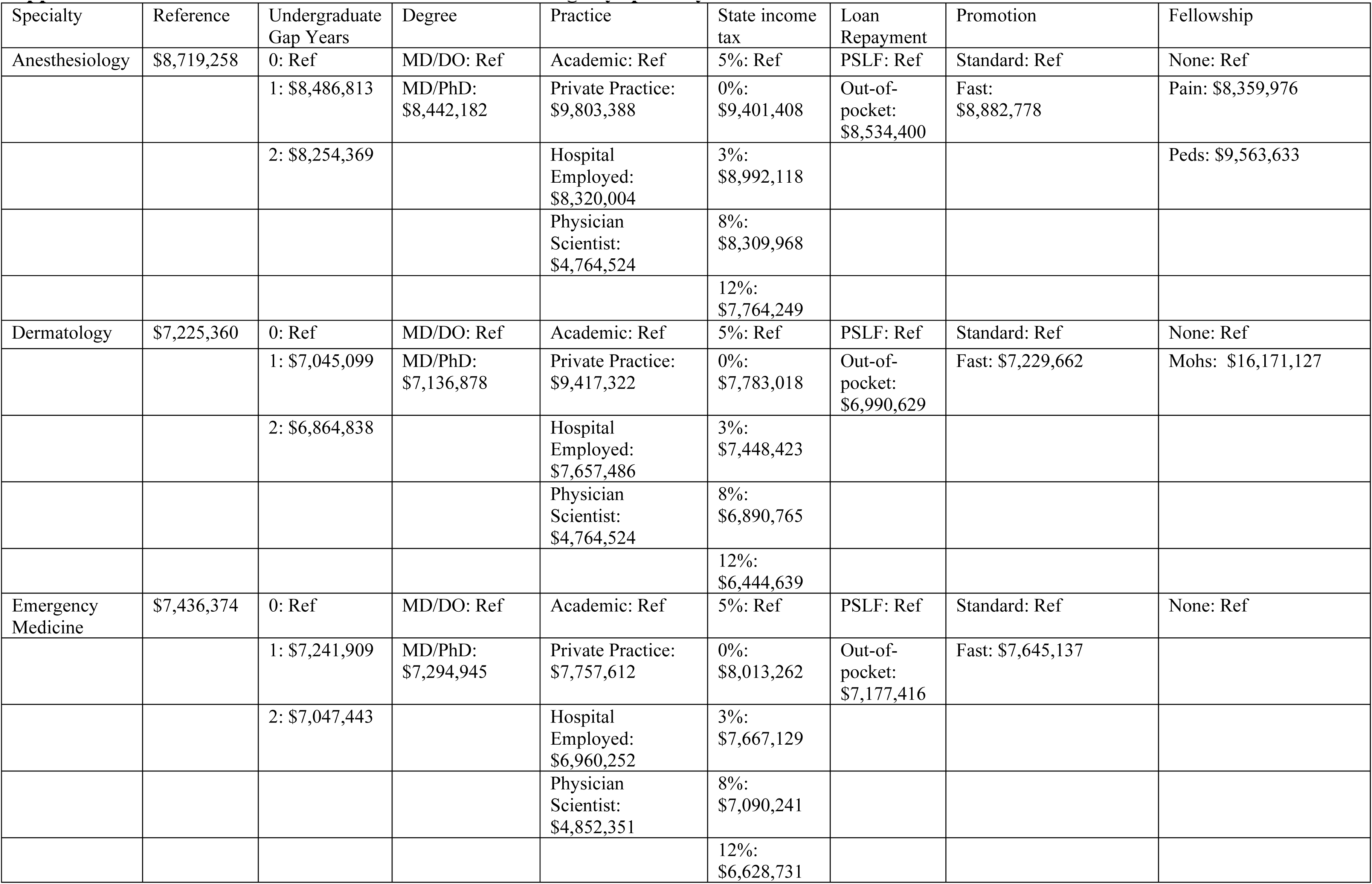
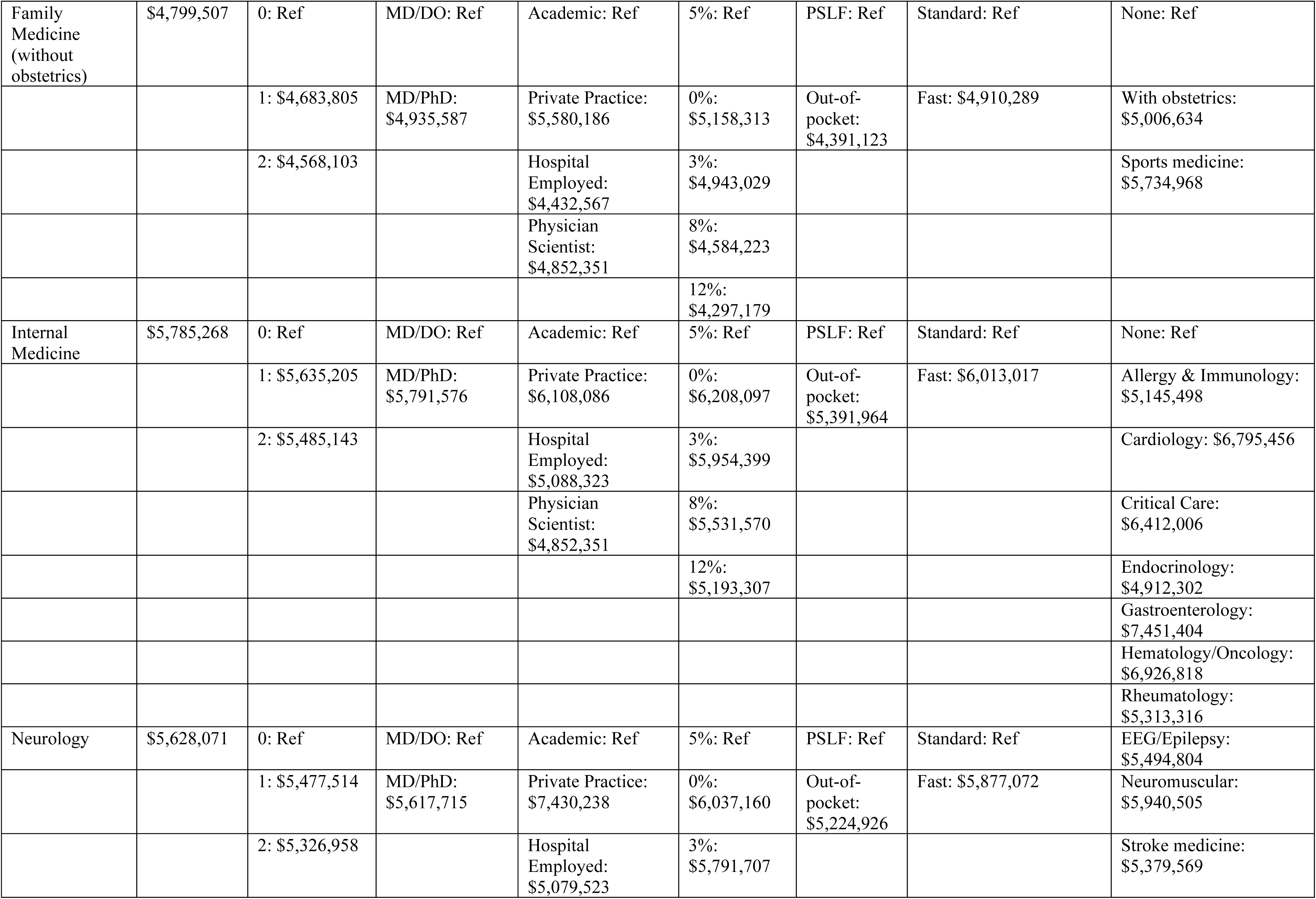
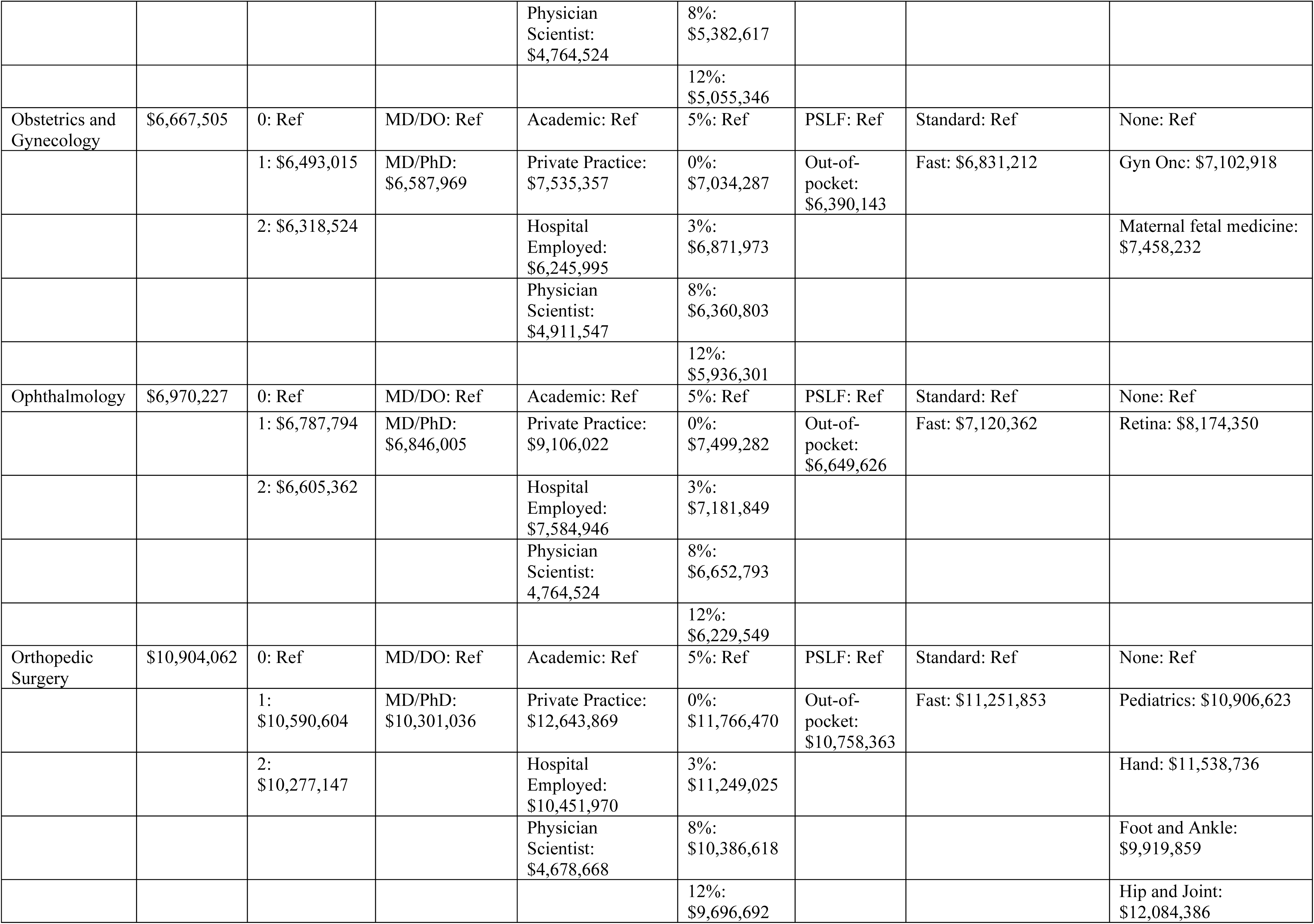
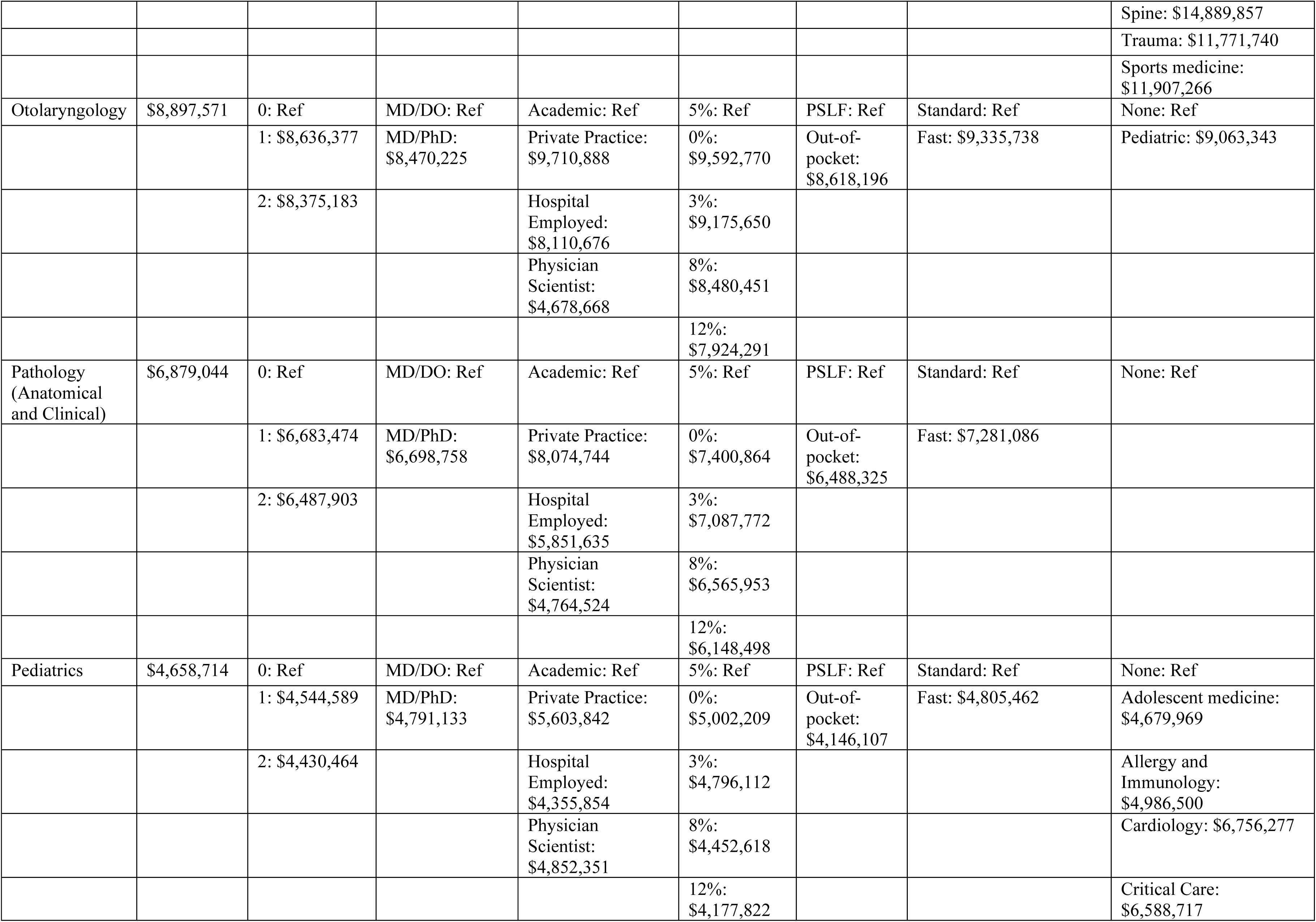
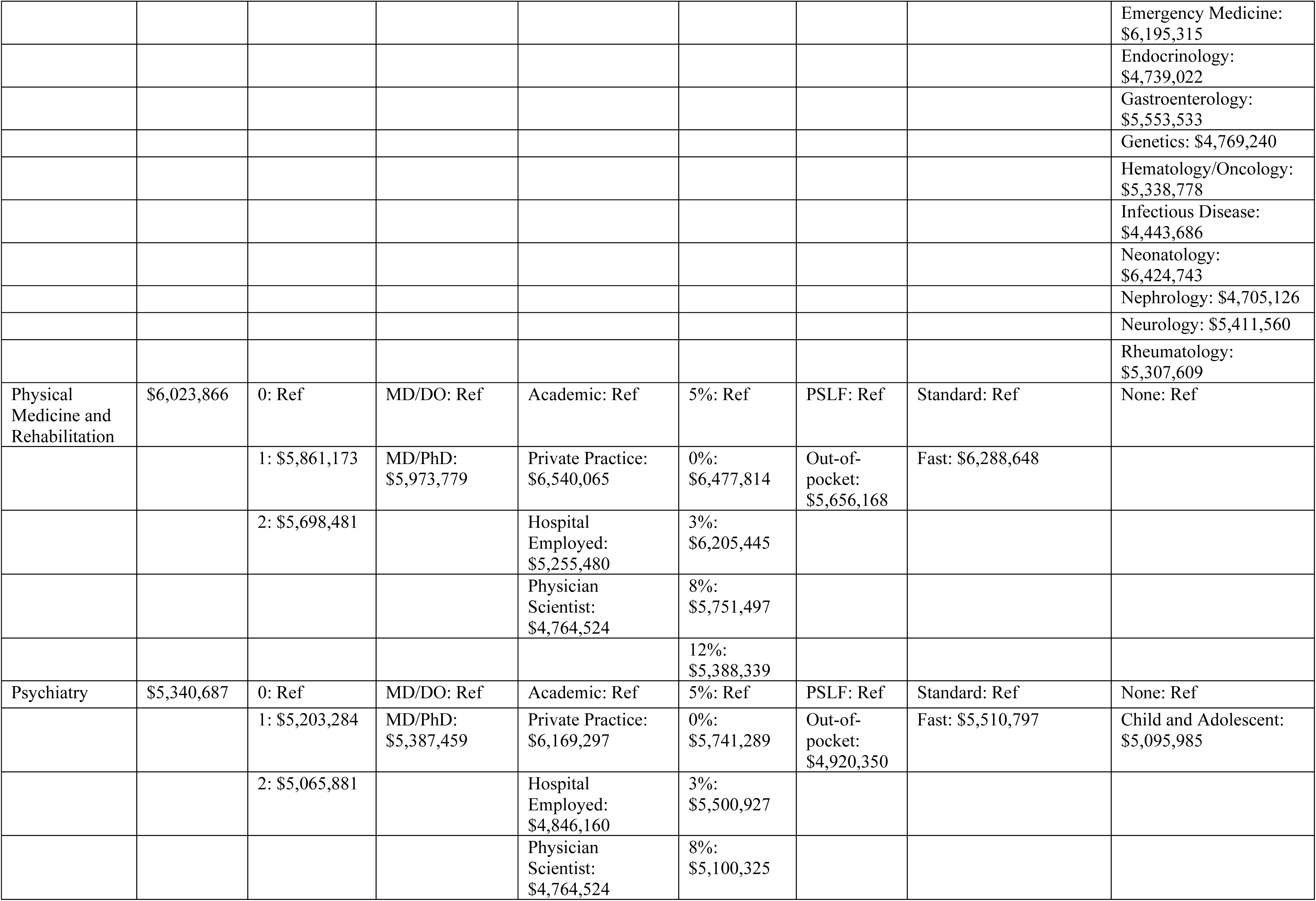
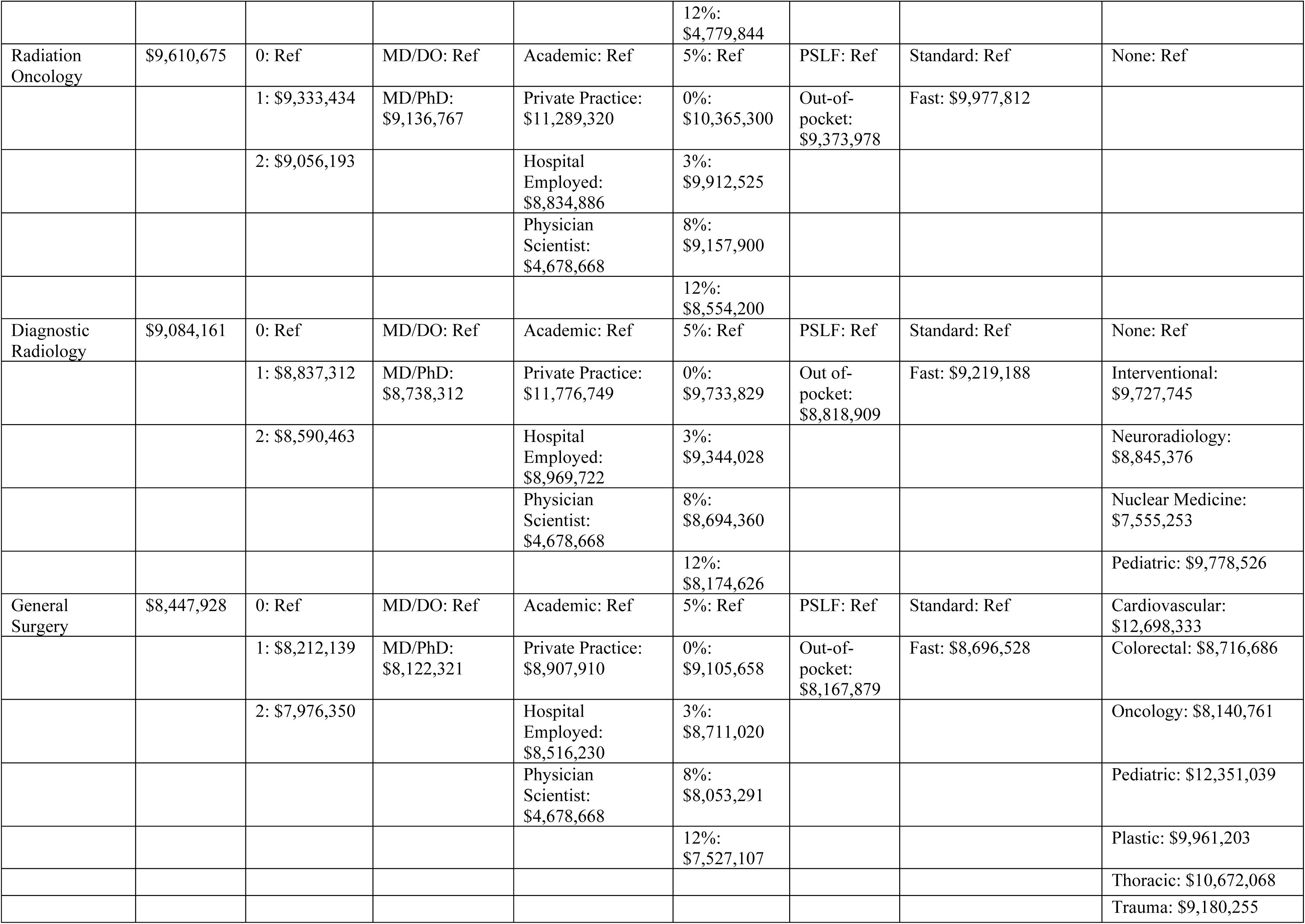
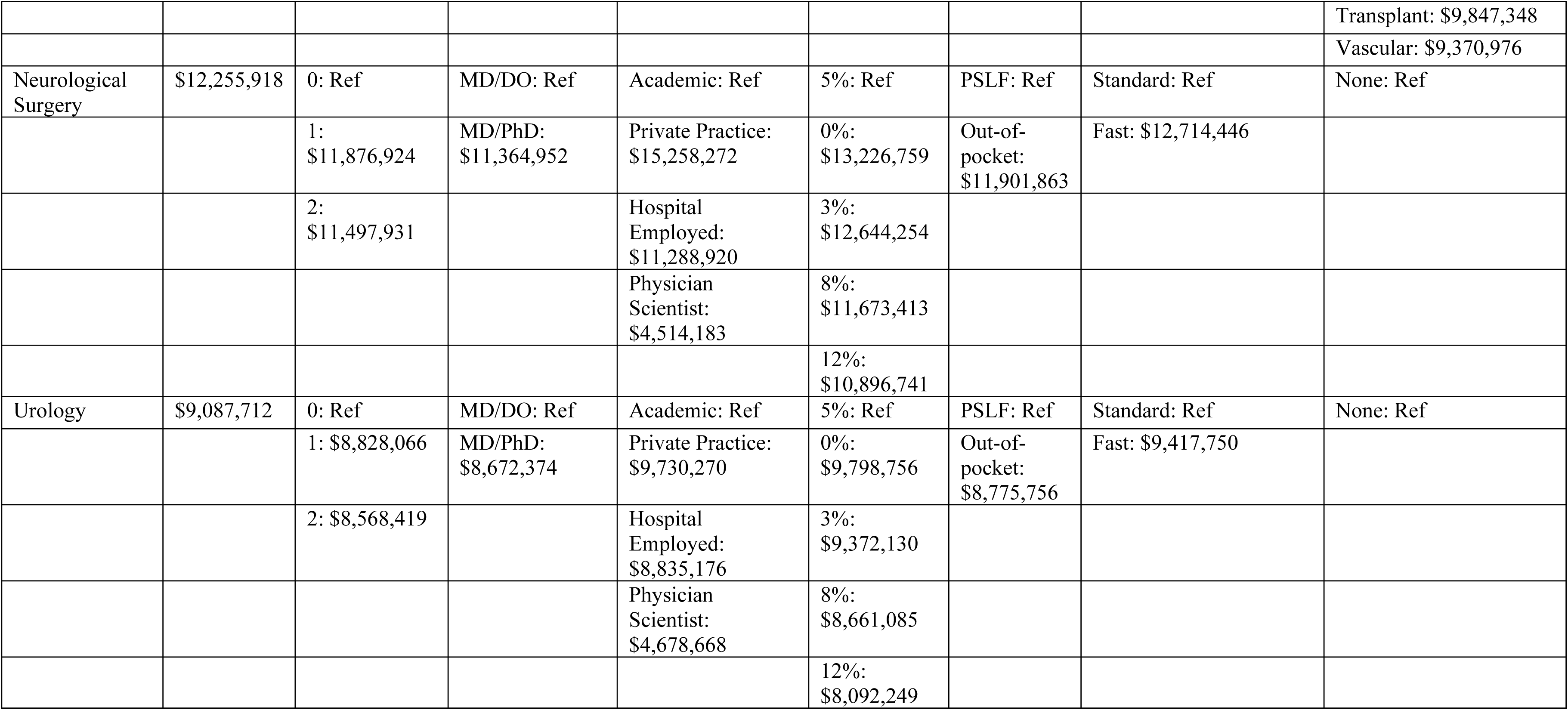
Cumulative lifetime median earnings by specialty.

**Supplemental Table 2.** Break-even age for the weighted mean of cumulative lifetime earnings across 19 selected specialties.

|  |  |  |
| --- | --- | --- |
| Group | Scenario | Break-even Age |
| All specialties | Reference level across all categories | 32 |
|  | Two gap years vs. zero gap years | 33 |
|  | MD-PhD degree vs. MD | 27 |
|  | Private practice vs. academia | 31 |
|  | Out-of-pocket loan repayment vs. PSLF | 33 |
|  | Accelerated academic promotion timeline (4 vs. 7 years each) | 32 |
| Primary care (family medicine, internal medicine, and pediatrics) | Reference level across all categories | 32 |
|  | Two gap years vs. zero gap years | 33 |
|  | MD-PhD degree vs. MD | 27 |
|  | Private practice vs. academia | 31 |
|  | Out-of-pocket loan repayment vs. PSLF | 33 |
|  | Accelerated academic promotion timeline (4 vs. 7 years each) | 32 |
| All specialties other than primary care | Reference level across all categories | 32 |
|  | Two gap years vs. zero gap years | 33 |
|  | MD-PhD degree vs. MD | 27 |
|  | Private practice vs. academia | 31 |
|  | Out-of-pocket loan repayment vs. PSLF | 32 |
|  | Accelerated academic promotion timeline (4 vs. 7 years each) | 32 |

**Supplemental Table 3.** Comparison of Median Lifetime Earnings Under Probabilistic Sensitivity Analysis vs. Deterministic Base Case.

| Specialty | PSA Median (\$) | Deterministic (\$) | % Change |
| --- | --- | --- | --- |
| Orthopedic (Nonsurgical) | \$10,144,471 | \$9,317,021 | 8.9% |
| Ophthalmology | \$7,870,733 | \$7,531,361 | 4.5% |
| Orthopedic: Hip and Joint | \$13,288,458 | \$12,744,208 | 4.3% |
| Orthopedic: Sports Medicine | \$13,024,735 | \$12,549,981 | 3.8% |
| Orthopedic (General) | \$11,983,676 | \$11,593,640 | 3.4% |
| Internal Medicine: Cardiology - Invasive | \$7,241,878 | \$7,026,482 | 3.1% |
| Surgery: Plastic and Reconstruction | \$10,936,997 | \$10,611,890 | 3.1% |
| Orthopedic: Pediatric | \$11,890,209 | \$11,562,748 | 2.8% |
| Internal Medicine: Hematology/Oncology | \$7,763,643 | \$7,554,607 | 2.8% |
| Orthopedic: Trauma | \$12,750,971 | \$12,427,996 | 2.6% |
| Surgery: Neurological | \$13,193,328 | \$12,867,509 | 2.5% |
| Dermatology: Mohs Surgery | \$17,329,023 | \$16,929,454 | 2.4% |
| Surgery: Vascular | \$10,278,325 | \$10,045,100 | 2.3% |
| Internal Medicine: Cardiology - Noninvasive | \$7,493,933 | \$7,337,999 | 2.1% |
| Internal Medicine: Pulmonary Medicine: General and Critical Care | \$8,351,282 | \$8,186,890 | 2.0% |
| Otorhinolaryngology | \$9,732,134 | \$9,553,771 | 1.9% |
| Internal Medicine: Gastroenterology | \$8,236,547 | \$8,091,178 | 1.8% |
| Internal Medicine: Pulmonary Medicine - Critical Care | \$6,619,899 | \$6,505,066 | 1.8% |
| Ophthalmology: Retina | \$9,983,317 | \$9,813,560 | 1.7% |
| Internal Medicine: Cardiology - Invasive-Interventional | \$8,866,946 | \$8,716,616 | 1.7% |
| Orthopedic: Hand | \$12,372,779 | \$12,167,932 | 1.7% |
| Internal Medicine: Cardiology - Electrophysiology | \$9,337,032 | \$9,182,903 | 1.7% |
| Internal Medicine: Allergy & Immunology | \$5,846,416 | \$5,751,040 | 1.7% |
| OB/GYN: Gynecological Oncology | \$9,083,207 | \$8,958,122 | 1.4% |
| Surgery: Colon and Rectal | \$9,479,726 | \$9,352,274 | 1.4% |
| Internal Medicine: Critical Care | \$7,158,349 | \$7,064,002 | 1.3% |

| Specialty | PSA Median (\$) | Deterministic (\$) | % Change |
| --- | --- | --- | --- |
| Dermatology | \$8,007,044 | \$7,901,730 | 1.3% |
| Surgery: Trauma | \$9,954,930 | \$9,830,634 | 1.3% |
| Surgery: Cardiovascular | \$13,393,675 | \$13,242,103 | 1.1% |
| Anesthesiology: Pain | \$9,137,346 | \$9,037,100 | 1.1% |
| Internal Medicine: Rheumatology | \$5,820,961 | \$5,759,685 | 1.1% |
| Surgery: Oncology | \$8,846,811 | \$8,754,454 | 1.1% |
| Surgery: Transplant | \$10,586,053 | \$10,476,022 | 1.1% |
| Orthopedic: Foot and Ankle | \$10,692,106 | \$10,583,401 | 1.0% |
| Psychiatry: General | \$6,031,687 | \$5,972,747 | 1.0% |
| Internal Medicine: Hospice | \$5,751,434 | \$5,695,233 | 1.0% |
| Family Medicine: Sports Medicine | \$6,397,403 | \$6,336,331 | 1.0% |
| Psychiatry: Child and Adolescent | \$6,321,639 | \$6,262,407 | 0.9% |
| Internal Medicine: Oncology (only) | \$6,941,811 | \$6,877,421 | 0.9% |
| OB/GYN: Maternal and Fetal Medicine | \$9,441,146 | \$9,353,574 | 0.9% |
| Radiology: Interventional | \$10,482,343 | \$10,385,278 | 0.9% |
| Otorhinolaryngology: Pediatric | \$9,798,231 | \$9,714,265 | 0.9% |
| Internal Medicine: Pulmonary Medicine - General | \$6,464,199 | \$6,409,170 | 0.9% |
| Pediatrics (General) | \$5,280,666 | \$5,236,180 | 0.8% |
| Urology | \$9,831,960 | \$9,749,730 | 0.8% |
| Pediatrics: Emergency Medicine | \$6,467,085 | \$6,413,996 | 0.8% |
| Pediatrics: Neonatal Medicine | \$6,769,380 | \$6,714,935 | 0.8% |
| Internal Medicine: Hospitalist | \$7,129,646 | \$7,072,465 | 0.8% |
| Surgery: General | \$9,175,427 | \$9,104,249 | 0.8% |
| Pathology: Anatomic and Clinical | \$7,497,317 | \$7,439,223 | 0.8% |
| Physical Medicine and Rehabilitation | \$6,673,737 | \$6,622,791 | 0.8% |
| Pediatrics: Allergy/Immunology | \$5,519,786 | \$5,478,992 | 0.7% |
| OB/GYN (General) | \$7,383,420 | \$7,329,741 | 0.7% |
| Pediatrics: Critical Care | \$6,901,292 | \$6,851,700 | 0.7% |

| Specialty | PSA Median (\$) | Deterministic (\$) | % Change |
| --- | --- | --- | --- |
| Internal Medicine: Endocrinology | \$5,565,491 | \$5,526,311 | 0.7% |
| Neurology: Neuromuscular | \$6,346,753 | \$6,302,240 | 0.7% |
| Anesthesiology | \$9,478,834 | \$9,415,770 | 0.7% |
| Internal Medicine: Gastroenterology - Hepatology | \$8,345,753 | \$8,291,431 | 0.7% |
| Internal Medicine: Nephrology | \$6,601,350 | \$6,558,545 | 0.7% |
| Surgery: Thoracic | \$11,394,943 | \$11,321,067 | 0.7% |
| Radiology: Pediatric | \$10,508,194 | \$10,440,599 | 0.6% |
| Emergency Medicine | \$8,172,064 | \$8,121,433 | 0.6% |
| Internal Medicine (General) | \$6,272,203 | \$6,234,933 | 0.6% |
| Neurology | \$6,104,177 | \$6,069,282 | 0.6% |
| OB/GYN: Gynecology (only) | \$7,413,977 | \$7,373,105 | 0.6% |
| Radiation Oncology | \$10,341,117 | \$10,284,504 | 0.6% |
| Pathology: Anatomic | \$6,793,800 | \$6,757,899 | 0.5% |
| Pathology: Clinical | \$6,544,616 | \$6,511,176 | 0.5% |
| Pediatrics: Gastroenterology | \$6,158,320 | \$6,127,959 | 0.5% |
| Pediatrics: Child Development | \$4,747,543 | \$4,724,667 | 0.5% |
| Pediatrics: Cardiology | \$7,412,326 | \$7,378,216 | 0.5% |
| Internal Medicine: General | \$6,262,276 | \$6,234,933 | 0.4% |
| Surgery: Pediatric | \$13,001,728 | \$12,945,454 | 0.4% |
| Pediatrics: Hematology/Oncology | \$5,785,790 | \$5,761,183 | 0.4% |
| Pediatrics: Hospitalist | \$6,149,553 | \$6,124,024 | 0.4% |
| Pediatrics: Rheumatology | \$5,752,846 | \$5,729,569 | 0.4% |
| Family Medicine (without OB) | \$5,468,420 | \$5,447,023 | 0.4% |
| Pediatrics: Infectious Disease | \$4,977,651 | \$4,958,851 | 0.4% |
| Pediatrics: Nephrology | \$5,240,018 | \$5,221,114 | 0.4% |
| Radiology: Nuclear Medicine | \$8,249,045 | \$8,219,372 | 0.4% |
| Pediatrics: Neurology | \$6,031,417 | \$6,010,369 | 0.4% |
| Family Medicine (with OB) | \$5,677,367 | \$5,658,689 | 0.3% |

| Specialty | PSA Median (\$) | Deterministic (\$) | % Change |
| --- | --- | --- | --- |
| Internal Medicine: Infectious Disease | \$5,120,871 | \$5,104,655 | 0.3% |
| Anesthesiology: Peds | \$10,271,040 | \$10,239,786 | 0.3% |
| Neurology: Epilepsy/EEG | \$6,093,017 | \$6,074,704 | 0.3% |
| Internal Medicine: Geriatrics | \$5,663,204 | \$5,647,565 | 0.3% |
| Pediatrics: Endocrinology | \$5,253,723 | \$5,239,378 | 0.3% |
| Neurology: Stroke Medicine | \$5,996,495 | \$5,980,825 | 0.3% |
| Pediatrics: Genetics | \$5,301,479 | \$5,289,012 | 0.2% |
| Radiology: Neurological | \$9,514,586 | \$9,495,674 | 0.2% |
| Radiology: Diagnostic | \$9,037,292 | \$9,022,807 | 0.2% |
| Pediatrics: Adolescent Medicine | \$5,191,107 | \$5,186,129 | 0.1% |
| Pathology: Surgical | \$5,400,054 | \$5,395,487 | 0.1% |
| Pediatrics: Pulmonology | \$5,436,201 | \$5,434,147 | 0.0% |

